# Vaporized H_2_O_2_ decontamination against surrogate viruses for the reuse of N95 respirators in the COVID-19 emergency

**DOI:** 10.1101/2020.06.25.20140269

**Authors:** Ebru Oral, Keith K. Wannomae, Rachel L. Connolly, Joseph A. Gardecki, Hui Min Leung, Orhun K. Muratoglu, John Durkin, Ralph Jones, Cassidy Collins, Julian Gjore, Amanda Budzilowicz, Tareq Jaber

## Abstract

Decontamination of N95 respirators has become critical to alleviate PPE shortages for healthcare workers in the current COVID-19 emergency. The factors that are considered for the effective reuse of these masks are the fit, filter efficiency and decontamination/disinfection level both for SARS-CoV-2, which is the causative virus for COVID-19, and for other organisms of concern in the hospital environment such as Staphylococcus aureus or Clostridium difficile.

In its guidance entitled ‘Recommendations for Sponsors Requesting EUAs for Decontamination and Bioburden Reduction Systems for Surgical Masks and Respirators During the Coronavirus Disease 2019 (COVID19) Public Health Emergency’ (May 2020)[1], the FDA recommends a 6-log10 reduction in either the most resistant bacterial spores for the system or in a mycobacterium species to authorize the use of a decontamination method of N95 respirators for single or multiple users. While the goal is primarily inactivation against SARS-CoV-2, testing of decontamination methods against the virus may not always be available. For decontamination methods considered for only single users, the recommendation is a 6-log10 reduction in the infective virus concentration of 3 non-enveloped viruses or in the concentration of two Gram (+) and two Gram (-) bacteria. Based on these recommendations, we explored the efficacy of vaporized H2O2 (VHP) treatment of N95 respirators against surrogate viruses covering a wide range of disinfection resistance for emergency decontamination and reuse to alleviate PPE shortages for healthcare workers in the COVID-19 emergency.

## Background

In its guidance entitled “Recommendations for Sponsors Requesting EUAs for Decontamination and Bioburden Reduction Systems for Surgical Masks and Respirators During the Coronavirus Disease 2019 (COVID19) Public Health Emergency” (May 2020)[1], the FDA recommends a 6-log_10_ reduction in either the most resistant bacterial spores for the system or in a mycobacterium species to authorize the use of a decontamination method of N95 respirators for single or multiple users. While the goal is primarily inactivation against SARS-CoV-2, testing of decontamination methods against the virus may not always be available. For decontamination methods considered for only single users, the recommendation is a 6-log_10_ reduction in the infective virus concentration of 3 non-enveloped viruses or in the concentration of two Gram (+) and two Gram (-) bacteria. Based on these recommendations, we explored the efficacy of vaporized H_2_O_2_ (VHP) treatment of N95 respirators against surrogate viruses covering a wide range of disinfection resistance for emergency decontamination and reuse to alleviate PPE shortages for healthcare workers in the COVID-19 emergency.

## Methods

### Sample preparation

N95 respirators (3M, 1860S) were cut into five equal pieces. Some of these pieces were spiked from the outer side of the mask while others were spiked from the inner side of the mask. Mask straps were also spiked separately. Each piece was spiked with one type of virus.

### Viruses

*Porcine Parvovirus (PPV), Bovine viral diarrhea virus (BVDV), Feline Calcivirus (FCV), Herpes Simplex Virus (HSV) and Influenza A Virus (InfA)*

### Preparation of virus stocks

Virus-infected cells, cultivated in cell culture medium containing 0–10 % (v/v) fetal bovine serum, were either frozen or thawed once prior to virus harvest. The cell debris was sedimented by centrifugation and filtration of the virus-containing cell culture supernatant through a sterile filter. The virus stock solution was stored at ≤ −60 °C in aliquots until use. The titer of the virus stock was determined according to the Spearman-Kärber method [2,3].

### Viraltitration

To determine the virus titers of the stock solution, serial three-fold dilutions were prepared with cell culture medium at a non-cytotoxic dilution. Then, 100 μL aliquots of each dilution were added to 8 wells of a 96-well-microtiter plate with indicator cells (in 100 μL cell culture medium per well). The cells were cultivated at 36.5 ± 1.0°C under 5.0 ± 1% CO_2_ in a humidified atmosphere. After a specified cultivation period, the microtiter plates were inspected microscopically for virus-induced changes in cell morphology. The viral titers were determined according to the Spearman-Kärber method.

The detection limit of a sample analyzed for the viral load is defined by the total volume incubated with the indicator cells. To improve the detection limit, a large volume of the sample was analyzed (large volume plating). Briefly, 200 μL of the diluted sample was added to a predefined number of wells containing the indicator cells in 100 μL cell culture medium. The cells were cultivated for a specified incubation period. Then they were inspected microscopically for virus-induced changes in cell morphology. The indicator cells used for BVDV, PPV, HSV-1, FCV, and InfA were BT (Taurus turbinate cell line), StNeb (Swine testis), Vero76 (*C. aethiops*, African green monkey kidney cells), CRFK (*Felis catus*, kidney cortex cells) and MDCK (*Canis familaris*, canine kidney cells), respectively. The corresponding growth media were Complete MEM for CRFK, MDCK and StNeb cells, Complete RPMI for Vero76 cells, and Complete DMEM for BT cells.

### Viral sample preparation and spiking

The face piece or straps of several respirators were cut into equal-size pieces (n=5). Spiking at three different locations of the N95 masks were tested, namely the outer surface, the inner surface, and the straps. All spiking experiments were performed with a known concentration of virus at 5% spike ratio. Each piece was spiked with one type of virus. Three different viruses were used to challenge these masks. This process was performed at room temperature. The spiked material was placed in a sterile glass container, then left to dry in a biosafety cabinet for at least 1 hour.

### Viral inactivation by vaporized hydrogen peroxide (VHP) decontamination

The VHP process were performed inside a Class II Type A2 biological safety cabinet by B & V Testing (Needham Heights, MA), a certified vendor for the VHP system (ARD1000®, Steris, Mentor, OH). The hydrogen peroxide concentration exposure was 452 ± 35 ppm for 2.5 hours followed by aeration for approximately 4.5 hrs. Pieces of the spiked respirators were suspended across twine in a biosafety cabinet. Biological indicators (Steel spore discs of *Geobacillus stearothermophilus*, 1.7 × 10^6^, TTX-06, Crosstex) and chemical indicator strips (4-inch wide, GPS250-R, SPS Medical, Rush, NY), were placed in five different locations in the biosafety cabinet. After decontamination and aeration, both control and treated respirator pieces were each placed in 20mL of cell/virus specific media and on an orbital shaker for at least 10 minutes to evenly disperse media across the respirator. An aliquot was removed, diluted to a non-cytotoxic (non-interfering) dilution using cell culture media and titrated accordingly. A media control was used for comparison of virus titer. Media control was performed by spiking the same amount of virus into cell/virus specific media and left for the same duration as the sample.

### Recovery and viral load assessment

The virus was considered to be recovered from the sample if a virus titer obtained from the sample was within ±1 log_10_ of the positive control. If the difference in virus titer was greater than 1 log_10_ from the titer of the positive control, the virus recovery was considered incomplete. There was no significant interference caused by the test sample if the reduction of the virus titer determined in the non-cytotoxic dilution of the test sample compared to the virus titer determined in cell culture medium was <0.5 log_10_ or within the range of the 95% confidence limit. If the difference in virus titer was > 1.0 log_10_, there was clear interference of the test sample. If the difference in virus titer was > 0.5 and ≤ 1.0 log_10_, the path forward was evaluated based on the dilution factor, the quality of the cells, and the overall CPE versus controls.

## Results

Chemical indicators used at five different locations in the biosafety cabinet all changed color, indicating correct hydrogen peroxide exposure. Bacterial spore strips used to validate the decontamination procedure tested negative after 7 days of incubation.

The stock titers used to spike the respirators were different for the five viruses (Table 1). For PPV and BVDV, the virus concentration was maintained (<0.5 log_10_ from stock titer) when spiked and recovered from all respirator surfaces. For FCV and HSV, the virus concentration was maintained on the inner and outer surfaces but decreased (>0.5 log_10_ from stock titer) when recovered from the straps. In contrast, the infective virus concentration decreased for InfA (<0.5 log_10_) on all respirator surfaces.

**Table 1.**
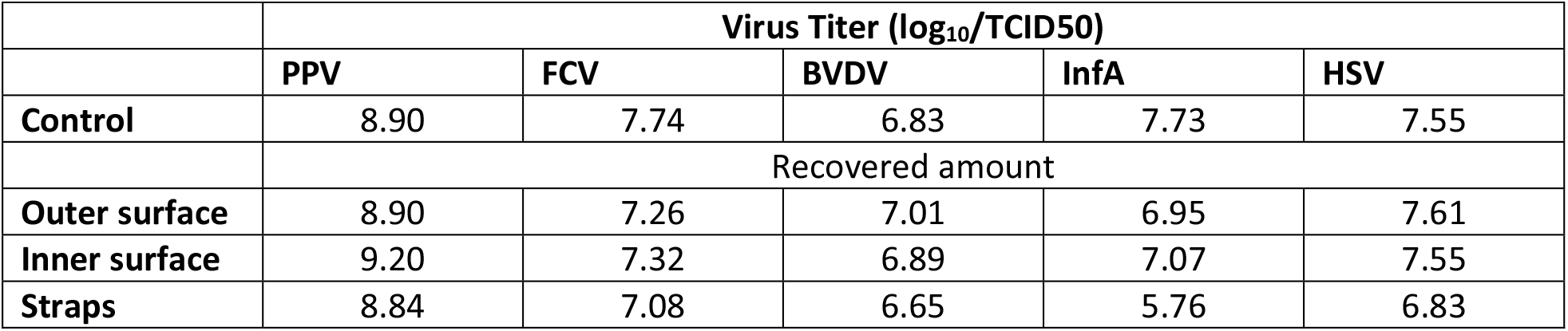
Stock titers and recovered amount of the different viruses from respirator surfaces.

|  |  |  |  |  |  |
| --- | --- | --- | --- | --- | --- |
| | Virus Titer ($\log_{10}$/TCID<sub>50</sub>) | | | | |
|  | PPV | FCV | BVDV | InfA | HSV |
| **Control** | 8.90 | 7.74 | 6.83 | 7.73 | 7.55 |
|  | Recovered amount |  |  |  |  |
| **Outer surface** | 8.90 | 7.26 | 7.01 | 6.95 | 7.61 |
| **Inner surface** | 9.20 | 7.32 | 6.89 | 7.07 | 7.55 |
| **Straps** | 8.84 | 7.08 | 6.65 | 5.76 | 6.83 |

For respirator samples spiked on the outer surfaces (Table 2), live virus was detected only on one out of five of the PPV-spiked samples after decontamination. For BVDV, for which the concentration of virus in the hold sample was also not detectable, a determination of the log-reduction due to the decontamination could not be made on the outer surfaces of the respirators. In contrast, there was at least 3.4-log_10_ reduction for all other viruses and at least a 6.0-log_10_ reduction for PPV after VHP decontamination of the outer surfaces of the spiked respirators.

**Table 2.** Concentration of infectious virus for untreated and decontaminated samples. The spiking was performed on the outer surface of the respirators. Green shading indicates detection of live virus plaque.

| *PPV (Parvoviridae)* |  |  |  |
| --- | --- | --- | --- |
| Sample | Viral load, PFU/sample | Viral load ($\log_{10}$) | Reduction ($\log_{10}$) |
| Control – viral inoculation => hold at RT | $7.41 \times 10^7$ | 7.87 | |
| Test – viral inoculation => decontamination | $\leq 0.79 \times 10^2$ | $\leq 1.90$ | **$\geq 5.97$** |
| | $0.28 \times 10^2$ | 1.44 | **6.43** |
| *BVDV (Flaviviridae)* |  |  |  |
| Control – viral inoculation => hold at RT | $\leq 2.63 \times 10^4$ | $\leq 4.42$ | |
| Test – viral inoculation => decontamination | $\leq 4.07 \times 10^2$ | $\leq 2.61$ | **Not detectable** |
| | $\leq 4.07 \times 10^2$ | $\leq 2.61$ | **Not detectable** |
| *FCV (Calciviridae)* |  |  |  |
| Control – viral inoculation => hold at RT | $8.51 \times 10^5$ | 5.93 | |
| Test – viral inoculation => decontamination | $\leq 0.79 \times 10^2$ | $\leq 1.90$ | **$\geq 4.03$** |
| | $\leq 0.79 \times 10^2$ | $\leq 1.90$ | **$\geq 4.03$** |
| *InfA (Orthomyxoviridae)* |  |  |  |
| Control – viral inoculation => hold at RT | $4.07 \times 10^6$ | 6.61 | |
| Test – viral inoculation => decontamination | $\leq 0.79 \times 10^2$ | $\leq 1.90$ | **$\geq 4.31$** |
| | $\leq 0.79 \times 10^2$ | $\leq 1.90$ | **$\geq 4.31$** |
| *HSV (Herpesviridae)* |  |  |  |
| Control – viral inoculation => hold at RT | $2.09 \times 10^5$ | 5.32 | |
| Test – viral inoculation => decontamination | $\leq 0.79 \times 10^2$ | $\leq 1.90$ | **$\geq 3.42$** |
| | $\leq 0.79 \times 10^2$ | $\leq 1.90$ | **$\geq 3.42$** |

For respirator samples spiked on the inner surfaces (Table 3), live virus was detected on both of the PPV-spiked samples and one of the FCV-spiked samples. For FCV, HSV and InfA, for which the concentration of virus in the hold sample was also not detectable, a determination of the log-reduction due to the decontamination could not be made on the inner surfaces of the respirators. There was at least a 2.4-log_10_ reduction for BVDV and at least a 5.6-log_10_ reduction for PPV after VHP decontamination of the inner surfaces of the spiked respirators.

**Table 3.** Concentration of infectious virus for untreated and decontaminated samples. The spiking was performed on the inner surface of the respirators.

|  |  |  |  |
| --- | --- | --- | --- |
| *PPV (Parvoviridae)* |  |  |  |
| Sample | Viral load, PFU/sample | Viral load ($\log_{10}$) | Reduction ($\log_{10}$) |
| Control – viral inoculation => hold at RT | $3.02 \times 10^8$ | 8.48 | |
| Test – viral inoculation => decontamination | $7.08 \times 10^2$ | 2.85 | **5.61** |
| | $0.28 \times 10^2$ | 1.44 | **7.02** |
| *BVDV (Flaviviridae)* |  |  |  |
| Control – viral inoculation => hold at RT | $9.12 \times 10^4$ | 4.96 | |
| Test – viral inoculation => decontamination | $\leq 4.07 \times 10^2$ | $\leq 2.61$ | **$\geq 2.35$** |
| | $\leq 4.07 \times 10^2$ | $\leq 2.61$ | **$\geq 2.35$** |
| *FCV (Calciviridae)* |  |  |  |
| Control – viral inoculation => hold at RT | $\leq 2.63 \times 10^3$ | $\leq 3.42$ | |
| Test – viral inoculation => decontamination | $2.19 \times 10^2$ | 2.34 | **Not detectable** |
| | $\leq 0.79 \times 10^2$ | $\leq 1.90$ | **Not detectable** |
| *InfA (Orthomyxoviridae)* |  |  |  |
| Control – viral inoculation => hold at RT | $\leq 2.63 \times 10^4$ | $\leq 4.42$ | |
| Test – viral inoculation => decontamination | $\leq 0.79 \times 10^2$ | $\leq 1.90$ | **Not detectable** |
| | $\leq 0.79 \times 10^2$ | $\leq 1.90$ | **Not detectable** |
| *HSV (Herpesviridae)* |  |  |  |
| Control – viral inoculation => hold at RT | $\leq 2.63 \times 10^4$ | $\leq 4.42$ | |
| Test – viral inoculation => decontamination | $\leq 0.79 \times 10^2$ | $\leq 1.90$ | **Not detectable** |
| | $\leq 0.79 \times 10^2$ | $\leq 1.90$ | **Not detectable** |

For respirator samples spiked on the straps (Table 3), no live virus plaques were detected for any samples after decontamination. For FCV, HSV, BVDV and InfA, for which the concentration of virus in the hold sample was also not detectable, a determination of the log-reduction due to the decontamination could not be made on the straps of the respirators. For PPV, there was at least a 6.3-log_10_ reduction after VHP decontamination of the straps of the spiked respirators.

## Discussion

In this study, one cycle of VHP sterilization (Steris ARD-100®) for the 3M 1860S N95 respirator was found to be effective in the inactivation of five different viruses with varying resistance to disinfection. A VHP system provided by the Battelle Memorial Institute (https://www.fda.gov/media/136529/download) and several low temperature sterilization systems by Steris (https://www.fda.gov/media/136843/download) with H_2_O_2_ decontamination cycles similar to those tested in this study, have obtained an Emergency Use Authorization for this application. These systems are approved for use for up to 20 and 10 decontamination cycles, respectively.

The concentration and time of hydrogen peroxide exposure tested here are designed for sterility assurance level of 10^−6^ (6-log_10_ reduction) with *Geobacillus stearothermophilus* spores. Process validation using these spores for the application/load was performed and verified with chemical indicators. Thus, H_2_O_2_ exposure under these conditions provided 6-log_10_ bioburden reduction of bacterial spores, which are the most resistant microorganisms. A ‘high level of disinfection’, also against less resistant common clinical pathogens such as Staphylococcal species and *C. difficile*, is expected as indicated in the FDA guidance entitled “Recommendations for Sponsors Requesting EUAs for Decontamination and Bioburden Reduction Systems for Surgical Masks and Respirators During the Coronavirus Disease 2019 (COVID19) Public Health Emergency” (May 2020). In this document, the FDA also recommends the demonstration of viricidal activity wherever possible (≥3-log).

The surrogate viruses used in this study were Influenza A; a single-stranded, enveloped RNA virus (80-120 nm), bovine viral diarrhea virus, a single stranded, enveloped RNA virus (40-50 nm), herpes simplex virus; a double-stranded, enveloped DNA virus (120-200 nm), feline calicivirus; a single stranded, non-enveloped RNA virus (35-39 nm) and porcine parvovirus; a single stranded, non-enveloped DNA virus (18-24 nm). In general, DNA viruses are more resistant to disinfection than RNA viruses, non-enveloped viruses are more resistant than enveloped viruses and smaller size viruses are more resistant than larger size viruses. Thus, these five viruses represent a spectrum of resistance towards disinfection. SARS-CoV-2, the causative virus of COVID19, against which decontamination of respirators is our focus, is a single stranded, enveloped RNA virus of 65-120 nm. Because of the viral properties of this virus, low-level disinfection is likely sufficient for its recommended inactivation (3-log reduction in the concentration of infective virus). Nevertheless, there is still little information about its persistence in different environments as well as its susceptibility to cleaning, disinfection and decontamination procedures [4,5].

The highest concentration that can be obtained before decontamination is dependent on how concentrated a stock solution can be for any virus and, also on the persistence of the virus on the given surface. Time-dependent effects are incorporated by comparing the final concentration of the samples to a ‘hold’ sample, which spends the same amount of time on the surface without the decontamination procedure. For the five viruses that we considered, infA showed a decrease in concentration simply due to the application on the respirator surface (Table 1; 0.78, 0.66 and 1.97 log_10_ on the outer surface, inner surface and straps, respectively). PPV showed a detectable but small decrease in concentration due to the hold time (1.03 log_10_, 0.72 log_10_ and 0.61 log_10_ decrease for the outer surface, inner surface and straps, respectively). However, all other viruses showed significant decreases in infective virus concentration due to the hold time alone (Tables 2, 3 and 4). In fact, in many cases, the there was no detectable infective virus concentration after the hold time; thus, the efficacy of the decontamination could not be quantitatively determined. In other cases, because the concentration of infective virus was low (3 or 4 log_10_), the log-reduction which could be shown due to decontamination was less than the recommended 6 log_10_ despite the lack of infective plaque forming units in culture. These are technical limitations associated with viral infectivity studies for the purpose of determining the efficacy of a decontamination method on respirator surfaces.

**Table 4.**
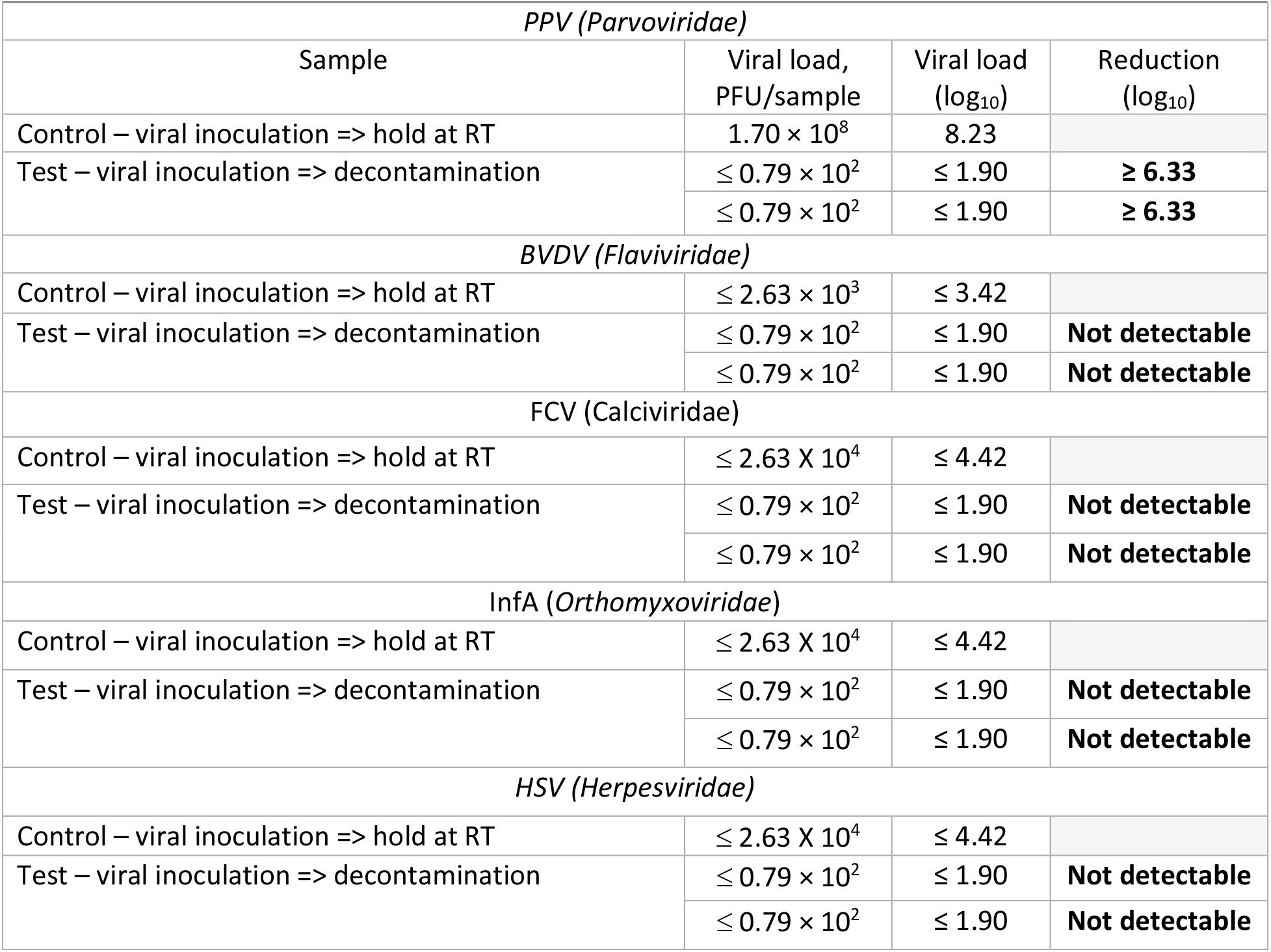
Concentration of infectious virus for untreated and decontaminated samples. The spiking was performed on the straps of the respirators.

|  |  |  |  |
| --- | --- | --- | --- |
| *PPV (Parvoviridae)* |  |  |  |
| Sample | Viral load, PFU/sample | Viral load ($\log_{10}$) | Reduction ($\log_{10}$) |
| Control – viral inoculation => hold at RT | $1.70 \times 10^8$ | 8.23 | |
| Test – viral inoculation => decontamination | $\leq 0.79 \times 10^2$ | $\leq 1.90$ | $\geq 6.33$ |
| | $\leq 0.79 \times 10^2$ | $\leq 1.90$ | $\geq 6.33$ |
| *BVDV (Flaviviridae)* |  |  |  |
| Control – viral inoculation => hold at RT | $\leq 2.63 \times 10^3$ | $\leq 3.42$ | |
| Test – viral inoculation => decontamination | $\leq 0.79 \times 10^2$ | $\leq 1.90$ | Not detectable |
| | $\leq 0.79 \times 10^2$ | $\leq 1.90$ | Not detectable |
| *FCV (Calciviridae)* |  |  |  |
| Control – viral inoculation => hold at RT | $\leq 2.63 \times 10^4$ | $\leq 4.42$ | |
| Test – viral inoculation => decontamination | $\leq 0.79 \times 10^2$ | $\leq 1.90$ | Not detectable |
| | $\leq 0.79 \times 10^2$ | $\leq 1.90$ | Not detectable |
| *InfA (Orthomyxoviridae)* |  |  |  |
| Control – viral inoculation => hold at RT | $\leq 2.63 \times 10^4$ | $\leq 4.42$ | |
| Test – viral inoculation => decontamination | $\leq 0.79 \times 10^2$ | $\leq 1.90$ | Not detectable |
| | $\leq 0.79 \times 10^2$ | $\leq 1.90$ | Not detectable |
| *HSV (Herpesviridae)* |  |  |  |
| Control – viral inoculation => hold at RT | $\leq 2.63 \times 10^4$ | $\leq 4.42$ | |
| Test – viral inoculation => decontamination | $\leq 0.79 \times 10^2$ | $\leq 1.90$ | Not detectable |
| | $\leq 0.79 \times 10^2$ | $\leq 1.90$ | Not detectable |

VHP decontamination of respirator surfaces spiked with PPV could be reliably quantified due to (1) high initial stock concentration and sustained concentration of infective units of PPV on surfaces; (2) high persistent concentration of infective units of PPV on the surfaces for a prolonged period of (hold) time; and (3) the detection of live infective units in culture after decontamination. VHP decontamination of respirator surfaces spiked with PPV ranged from 5.61 to 7.02 log_10_ reduction, showing a high level of disinfection/inactivation against this non-enveloped virus. We have previously shown inactivation of infective SARS-Cov-2 using VHP decontamination under similar conditions to those tested here [6], supporting the efficiency of this decontamination method against a wide range of pathogens.

We have also previously shown that the filter efficiency and fit for the 3M 1860S N95 respirator was not detrimentally affected by VHP decontamination for at least one cycle of decontamination [6]. The results of our study only consider bioburden reduction, one of the many considerations that healthcare facilities need to consider in selecting a decontamination method to utilize during emergency use of N95 respirators. The results presented here are not meant to constitute a stand-alone recommendation.

## Data Availability

The data is listed in the preprint and any further details are available upon contact of the authors.

